# Epidemiology and preclinical management of dog bites among humans in Wakiso and Kampala districts, Uganda: Implications for prevention of dog bites and rabies

**DOI:** 10.1101/2020.02.16.20023556

**Authors:** Stevens Kisaka, Fredrick E. Makumbi, Samuel Majalija, Alexander Bangirana, SM Thumbi

## Abstract

In rabies endemic areas, appropriate management of dog bites is critical in human rabies prevention. Victims must wash bite wounds for 15 minutes with soap, water and disinfectant immediately before seeking medical care. This study investigated the epidemiology of dog bites and determinants of compliance to these pre-clinical guidelines requirements among dog bite victims from high rabies-burden areas of Wakiso and Kampala, Uganda. An explanatory sequential mixed-methods study design was used. Quantitative data were collected from 376 dog-bite patients at two healthcare facilities. Qualitative data as also collected through 13 in-depth interviews with patients, healthcare workers, herbalists and veterinarians. Qualitative data were analyzed using a deductive thematic approach. Generalized lineal models were used to determine factors associated with compliance. Nearly half (190, 51%) of the patients were from Wakiso District and 293 (77.9%) had grade II wounds. Most of the wounds (171, 45.5%) were on the legs. Two-thirds of the bites occurred in public places. Only 70 (19%) of the bite patients had complied with pre-clinical guidelines. Nearly half of the patients had applied un-recommended substances such as herbs (47/193), antiseptics (46/193), “black stone” (25/193) and unknown creams (10/193) on the wounds. Factors negatively associated with compliance included: being aged 15 years or older, adjPR = 0.70 (0.47 - 0.92) and knowing the dog owner, adjPR=0.65 (0.36 - 0.93). However, attainment of secondary or higher education, adjPR= 1.76 (1.24 – 3.79), being in employment, adjPR = 1.48 (1.09 – 2.31), perception that the dog was sick, adjPR = 1.47 (1.02 – 2.72) and knowledge about the dog’s subsequent victim(s) adjPR=0.35 (0.17 - 0.70) were positively associated with compliance. High occurrence of dog bites in public places by free-roaming dogs suggests the need for promotion of responsible dog ownership. Additionally, targeted health education may be required to improve the low compliance to pre-clinical guidelines.

**Author summary:** Dog-mediate rabies is on the rise, especially in sub Saharan Africa. Though the disease is fatal upon exposure, it can be effectively prevented through appropriate post-exposure management. It is recommended that dog bite victims wash bite wounds for 15 minutes with soap, water and disinfectant immediately before seeking medical care. However, such pre-clinical recommendations are not usually followed in many societies, including in Uganda. There are numerous reports of victims not seeking or delaying to seek healthcare. Additionally, victims have been reported not to wash their wounds and applying traditional herbal concoctions before presenting at health facilities. Such divergence from the recommended standards has negative implications on the effectiveness of post-exposure prophylaxis that is given when victims report to health facilities. Our study investigated the epidemiology of dog bites and preclinical practices for the victims in the context of dog bite prevention and rabies prevention respectively. We call for targeted health education programs to improve pre-clinical behavior, regulation of herbalist activities and interventions that minimize human-dog interactions.

## Introduction

Rabies, a neglected tropical disease, is estimated to cause 59,000 human deaths, over 3.7 million disability-adjusted life years (DALYs) and USD 8.6 billion in economic losses worldwide annually [1]. Although the rabies virus can infect all mammals, over 99% of all human rabies cases are transmitted through dog bites [2]. Consequently, in addition to mass dog vaccination that breaks rabies transmission cycles, strategies for prevention of rabies in humans include prevention of dog bites and appropriate post-exposure treatment (PET) [2, 3].

World Health Organization (WHO) has developed guidelines for dog bite victims before presenting to a healthcare facility (preclinical management) as well as how the cases must be managed in the healthcare facility (clinical guidelines) [4]. These preclinical guidelines are summarized as: wash the bite wound with running water for 15 minutes; disinfect the wound with substances with capacity to kill the rabies virus (soap, disinfectant); and seek medical care immediately to receive post-exposure prophylaxis vaccines. Appropriate washing and disinfection of wounds can prevent one-third of rabies infections [3, 5]. Inadequate dog bite wound care has been associated with increased likelihood of PET failure and progression to rabies [5, 6].

Pre-clinical practices that deviate from recommendations have been reported to include not seeking medical care following dog-bites [7], delay in seeking treatment [3, 8, 9], lack of wound washing or treatment of wounds with chilies, salt, turmeric powder, lime, snuff powder, paste of leaves, acid and ash provided by traditional healers and magicians [10, 11]. Non-compliance to the preclinical guidelines has been attributed to geographical, social, economic, cultural, organizational, dog and wound factors [12-15].

In Uganda, there are an estimated over 30,000 animal bites reported to healthcare facilities annually and the burden keeps on rising despite ongoing interventions like health education [16]. The country had approximately 486 suspected human rabies deaths between 2001 and 2015 [17] although some authors have estimated the per capita annual death rate from rabies to be at 0.39/100,000 [18]. Despite such a high burden of bites and rabies, the reports of delays in seeking medical care and victims treating dog bite wounds with traditional herbal concoctions remain largely anecdotal. There is barely any published data on pre-clinical management of dog-bites in Uganda. In this study, we investigated the epidemiology and preclinical management of dog bites in Wakiso and Kampala districts, Uganda. We include data on circumstances of dog bites and what influences people’s responses dog-bites with a view of identifying opportunities for prevention of dog bites and rabies.

## Methods

### Study design and area

We used an explanatory sequential study with a mixed methods approach. This included collection and analysis of quantitative data followed by collection and analysis of qualitative data. The study was conducted in two referral healthcare facilities; Mulago National Referral Hospital (Kampala City Authority) and Entebbe General Referral Hospital (Wakiso district) in Uganda. Both facilities routinely provide dog bite post-exposure treatment in the two rabies endemic districts. Approximately 14% and 8% of households own an average number of 1.9 and 1.7 dogs per household in Wakiso and Kampala respectively [19]. From the perspective of interventions, the districts have the highest number of dogs vaccinated against rabies [20]. Details of the study sites are shown in **S1 Fig**.

### Study population and data collection

#### Quantitative data

All patients presenting with dog bites at the two study health facilities for first-time PET between April 2019 and October 2019 were enrolled upon providing informed consent to enroll in the study. Based on severity of the wounds, patients were classified in one of 3 categories: Category I (unbroken skin); Category II (superficial scratches without bleeding) and Category III (bites / scratches which penetrate the skin with bleeding). Patients with category I bite exposure (44/420, 11%) who were assessed as not requiring PET, were excluded from the study. Quantitative data including pre-clinical practices, socio-demographic factors, patient and biting dog factors and circumstances surrounding the bite incidence were collected as shown in **S1 Table**. All data collection tools were in English and translated into Luganda languages. Pre-testing of the questionnaires was completed on animal bite patients in Mukono Health Center IV, Mukono district, Uganda.

#### Qualitative data

In-depth interview (IDI) guides were used to collect qualitative data on dog bite circumstances and preclinical practices. In total, 13 IDIs were conducted with 7 patients, 3 health care workers, 1 herbalist and 2 local veterinary officers to understand different perspectives of health seeking by dog bite victims. Selection of patients for in-depth interviews was purposeful and based on their reported outstanding compliance or non-compliance to preclinical guidelines. This approach is generally used for collecting qualitative data [21]. IDIs were recorded using a digital audio recorder device (SONY ICD PX333 Digital Voice Recorder^®^). Key points brought up during interviews were also written down. Data saturation was determined to have been reached when no new or / and relevant information materialized from the additional interviews conducted.

### Data Management and analysis

#### Quantitative data

The outcome variable, “compliance” was recorded and categorized as “compliant” (if the patient had washed wound with water and soap in addition to seeking medical care within 24 hours) and “non-compliant” (if one of the former was missed or patient applied non-recommended substances to the wound).

Data were double-entered by independent data entrants into Epi-info version 7.1.4.0, cleaned and exported to STATA14 (StataCorp.; College Station, TX, USA) for analysis. Exploratory data analyses were conducted and generated descriptive statistics for the continuous and categorical variables. Median (IQR) for the continuous variables and percentages for categorical variables were computed. Compliance to the pre-clinical guidelines was coded as 1 if patient was “compliant” and 0 if “non-compliant”, to form a binary outcome variable. In the bivariate analyses, categorical variables of importance were tabulated against compliance. The association was based on chi-square and determined to be statistically significant if *p* < 0.05. In the multivariable analysis, prevalence ratios (PRs) were computed using a generalized linear model (GLM) analysis with Poisson family and a log link with robust standard errors. The model included variables with *p* < 0.25 at bivariate analysis or variables found to be potential or known to be associated with the outcome from the literature. Both the unadjusted and adjusted prevalence ratios and corresponding 95% confidence intervals are presented.

#### Qualitative data

Independent individuals transcribed the recorded data into written text. Each transcript was given to the respective data collectors to verify the transcripts. NVivo 11.4.1^®^ software (QSR International, 2017) was used to organize these data according to pre-set categories. The transcripts were reviewed to identify the information that is related to the pre-set categories and themes were developed. Under each theme, the information was inductively coded into sub-themes and then patterns identified to form the explanatory points of what is being observed. Key statements corresponding to the themes were presented together with quantitative findings.

### Ethical considerations

The study protocol was approved by University of Nairobi - Kenyatta National Hospital Ethics Review Committee (Kenya) REF: P687/09/2018; Mulago National Referral Hospital Research and Ethics Committee (Uganda) REF: MREC 1518; and the Uganda National Council of Science and Technology (Uganda) REF: SS4911. Written permission was obtained from hospitals before commencement of the study. Informed assent was obtained from participants as well as caretakers of minors prior to the study. For minors, assent was obtained after giving them an explanation of study purpose, procedure and their rights. All data were anonymized and handled confidentially.

## Results

The total number of dog-bite patients enrolled in the study was 376. Of these, 201(54%) were male, and the median (IQR) age was 18 (22.75)18 years. One hundred and ninety (50.5%) of the patients were from Wakiso district. Eleven percent of the bite-patients reported to own at least one dog while only 5.1% had ever been vaccinated against rabies. Nearly three-quarters (72%) had ever received some information about dogs and dog bites from sources including friends (46%), family (14%), school (10%), and books (4%). Some victims (8%) reported to have suffered dog-bites previously. A summary of the socio-demographic characteristics of the dog-bite patients, dog-ownership and sources of information on dog-bites for the study participants is provided in **Table 1**.

**Table 1:**
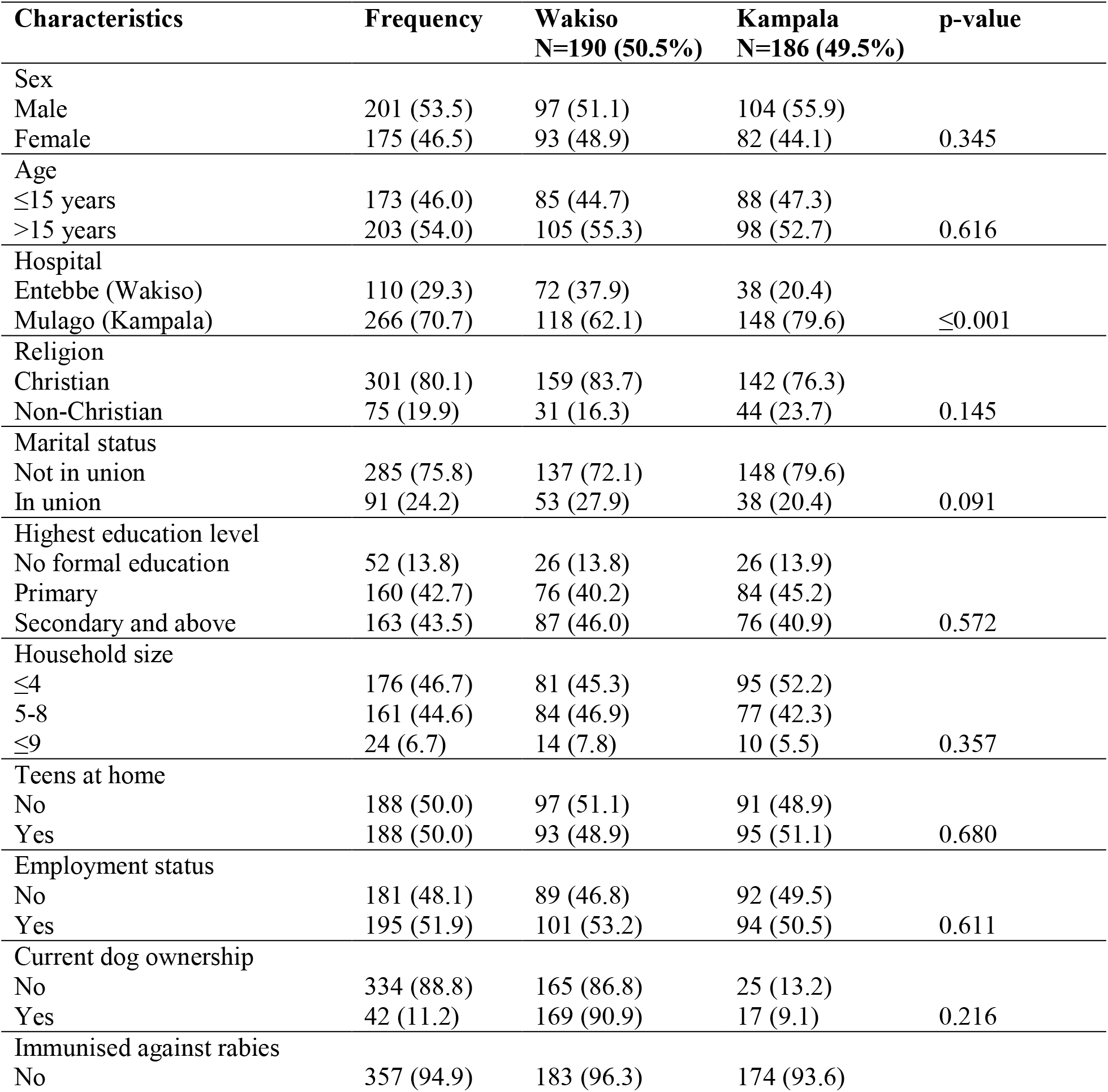
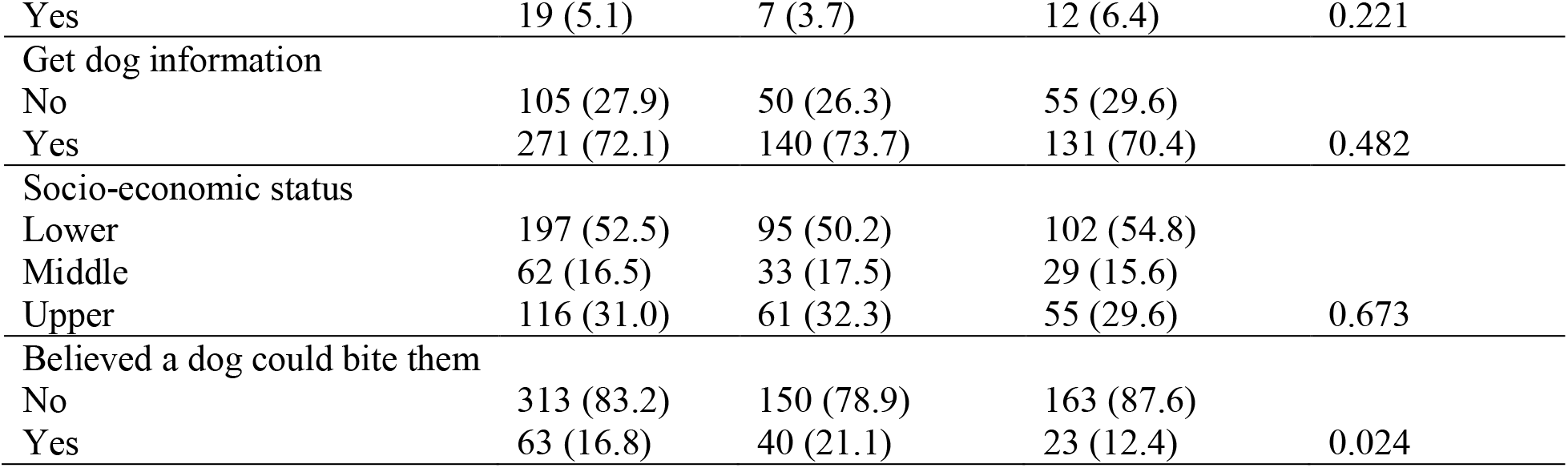
Characteristics of the 376 dog bite study participants stratified by district of bite event.

### Characteristics of dog bite injuries

Nearly two-thirds of the dog bite wounds (239/376, 63.7%) were single bites. Three-quarters (293/376, 77.9%) of the wounds were grade II and the rest were grade III. Forty-six percent of the bite patients had wounds on their legs, 14% on the head, 3% on the face and 3% several bite sites. The dog-bite distribution by body part and age of bite-patients are summarized in **Table 2**.

**Table 2:** Age-specific dog bite distribution by body part among the 376 participants

| Age (yrs) | Leg | Thigh | Arm | Abdomen | Back | Head | Face | Other | Combination | Total |
| --- | --- | --- | --- | --- | --- | --- | --- | --- | --- | --- |
| ≤15 years | 62 | 31 | 17 | 3 | 16 | 25 | 7 | 3 | 9 | 173 |
| Percentage | 35.8 | 17.9 | 9.8 | 1.7 | 9.3 | 14.5 | 4.1 | 1.7 | 5.2 | 100.0 |
| >15 years | 109 | 38 | 7 | 0 | 10 | 29 | 4 | 3 | 3 | 203 |
| Percentage | 53.7 | 18.7 | 3.5 | 0.0 | 4.9 | 14.3 | 1.9 | 1.5 | 1.5 | 100.0 |
| Total | 171 | 69 | 24 | 3 | 26 | 54 | 11 | 6 | 12 | 376 |
| Percentage | 45.5 | 18.4 | 6.4 | 0.8 | 6.9 | 14.4 | 2.9 | 1.6 | 3.2 | 100.0 |

### Characteristics of the biting dogs

Seventeen percent of the dog-bite patients had been bitten by their own dogs while 46% of the victims knew the owner of the dog that bit them. Nearly a third (30%) of the bite patients could identify the offending dog. Of these 113 biting dogs known to the victim, 21% had been vaccinated against rabies, 26% had not been vaccinated, and 53% were of unknown vaccination status. The victims described the dog as being male in 35% of the cases, 19% female and the rest were of unknown sex. The details on the characteristics of the biting dogs are presented in **S2 Table**.

### Circumstances of dog bites

Most of the dog bites (166/376, 44.2%) occurred in the afternoons (12 noon – 6pm) and the least (58/376, 15.4%) happened at night (7pm – 5am). Majority of the bites (339, 90%) were unprovoked and 137 (37%) of the bites occurred when the persons bitten were walking on the road. Nearly all the biting dogs (324, 86%) were unrestrained without a leash. **Table 3** summarizes data on circumstances surrounding the bites as reported by the bite patients.

**Table 3:** Circumstances of dog bite events among the 376 dog bite victims seeking post-exposure prophylaxis in the 2 selected hospitals in Uganda.

| Circumstances /contextual factor | Frequency (n) | Percentage (%) |
| --- | --- | --- |
| What time of day did the dog bite event happen? |  |  |
| Morning | 152 | 40.4 |
| Afternoon | 166 | 44.2 |
| Evening / night | 58 | 15.4 |
| Was it raining when the dog bite event happened? |  |  |
| No | 347 | 92.3 |
| Yes | 29 | 7.7 |
| If the dog bite happened at night, was there a visible moon? |  |  |
| No | 27 | 46.5 |
| Yes | 31 | 53.5 |
| Was the owner around when the bite happened? |  |  |
| No | 255 | 67.8 |
| Yes | 121 | 32.2 |
| Did victim previously know the biting dog? |  |  |
| No | 262 | 69.9 |
| Yes | 113 | 30.1 |
| Where did the event happen (place of event)? |  |  |
| Own home | 124 | 33.0 |
| Premises of person known to victim | 86 | 22.9 |
| Premises of person not known to victim | 4 | 1.1 |
| On the road | 137 | 36.4 |
| Other (e.g. market, classroom) | 25 | 6.6 |
| Was the victim in company of other people when dog bite occurred? |  |  |
| No | 211 | 56.1 |
| Yes | 165 | 43.9 |
| What was the victim doing just before the dog bite? |  |  |
| Walking | 209 | 55.6 |
| Seated | 46 | 12.2 |
| Chasing dog away | 8 | 2.1 |
| Feeding dog | 8 | 2.1 |
| Other | 105 | 27.9 |
| Was it the victim that approached the biting dog? |  |  |
| No | 37 | 9.8 |
| Yes | 339 | 90.2 |
| Was the biting dog on the leash? |  |  |
| No | 324 | 86.2 |
| Yes | 52 | 13.8 |
| Did the victim attempt to fend off the biting dog? |  |  |
| No | 218 | 58.0 |
| Yes | 158 | 42.0 |
| Did the victim think or feel that the dog intended to bite them? |  |  |
| No | 124 | 33.0 |
| Yes | 252 | 67.0 |
| Does the victim blame anyone for the bite? |  |  |
| No | 286 | 76.1 |
| Yes | 90 | 23.9 |
| What immediate action was taken against biting dog? |  |  |
| Chased it away | 91 | 24.1 |
| Killed it | 19 | 5.1 |
| Nothing | 177 | 47.1 |
| Ran away by itself | 83 | 22.1 |
| Other | 6 | 1.6 |

#### Circumstances of dog bites

##### Routine activities bringing dogs and humans into close proximity

Additional insights into dog bite circumstances were grouped as shown in **S3 Table**. A common view was that victims were bitten while undertaking everyday activities. Respondents spoke about holding something that drew the interest of the dog. Additionally, they talked about activities that brought dogs into close proximity with people as some explained:

> *“On my way back from the abattoir to buy meat, I didn’t know that there is a dog nearby, I only realized when it was holding onto my leg…… the dog continued biting me until a man came and hit it. By this time, it had even bitten my buttocks*.*” (Adult patient, female)*.
>
> *“We were playing with other children, running in circles in the compound. Our dog joined us and we ran with it. When I stopped, it jumped and bit me without warning*.*” (Patient, male child)*.

##### Disturbing dogs and threatening owner

However, some respondents explained that the biting dog had been deliberately disturbed either by themselves or by others. In addition, some thought that dogs also bit them when they acted in a way that threatened the dogs’ masters. Notably, such dogs had been on the loose in presence of strangers. One of the participants explained as follows;

> *“That Saturday morning, I went to visit my friend. We talked right in the compound, standing. However, when we laughed loud, I remember the dog barked. When we gave each other a ‘high-five’ and hugged, all I remember is the owner shouting at the dog to let go of my shirt. In the struggle, it bit me two times on the back and leg*.*” (Male adult patient)*.

##### Unusual behavior and protective tendencies

Some dog owners explained unusual behavior of the dog e.g. biting every living thing in the homestead, whether it posed a danger to it or not. They interpreted this as potentially rabid behavior. In addition, others were bitten by dogs protecting each other in a pack or with young ones as one explains;

> *“…. since our dog produced it does not want to interact with us. It no longer sits in front of the kitchen door as it used to do. I was with this boy in the kitchen and when I left to go to the house, he says he went behind the kitchen to see the dog and its babies. He said that is when it jumped and bit him on the shoulder. When I checked on the dog, it also wanted to bite me*.*” (caretaker / mother to a child patient)*.

##### Deviant handling practices

A number of respondents bitten by their own dogs explained circumstances that pointed to deviation from routine practices of handling dogs. They tended to inflict pain on the dogs during handling. In retaliation, the dogs bit them as one of them elaborates;

> *“Normally, I call them to follow me to their kernel and they do. But this time one of them refused and after taking in others, I went back and dragged it by the front leg. When it resisted, I lifted it and tried to push it into the house. This is when it bit my hand “(Adult male patient)*.

##### Seasons

For some, there were conditions like rain that caused interaction with the dog in open shelters. However, some described circumstances of having been bitten by dogs left unattended to, even without sharing shelter with them. On the other hand, some practitioners described bites as a seasonal issue linking them to late night activities especially during festive days as one explains;

> *“I get most of the people during big days* [festive] *like Christmas and Easter. This is when my house [serves as the care facility] is always full. Do you know why? People drink yet most of the dogs without owners also move at night. So they meet themselves and in most cases people harass these dogs first because they are scared of them. This is when they get bitten and come here for treatment*.*” (Herbalist attending to dog bite victims)*.

#### Immediate actions taken by dog bite victims

##### Reporting to local leaders and area veterinarians

When we inquired into what victims did immediately after the bite, the key actions included seeking medical care and legal action as summarized in **S3 Table**. Reporting to local authorities was quite common especially when victims wanted local leaders to put owners of the biting dog to task of owning up the responsibility of treating them. However, local veterinarians explained that some victims immediately call them because they know that it is their responsibility to ensure that dogs do not bite them. In other circumstances, the victims call veterinarians to seek treatment advice as one explains;

> *“They can call to be advised, others rush to the nearest health center and that is where they refer them to Entebbe hospital…*… *Many of them ask if my office has anti-rabies vaccines thinking such vaccines are kept with the area vet. They even get annoyed when I tell them I don’t have the vaccine*.*” (Local veterinarian)*.

##### Presenting to healthcare facility

Notably, there are some who immediately went to a healthcare facility. In comparison, some victims spent time regretting and filled with fear of bite consequences, especially death. Those that experienced this state related to the previous events that they had heard or witnessed in their lives as one narrates below;

> *“I cried, I just sat there and cried. I thought I was going to die. In our place* [of origin], *a dog bit a man and after 3 months he started barking like a dog, yes. All my thoughts ran to that man who died thinking like I was going to be like him. Besides, I was also in too much pain. You see this finger, I still feel paralysis and sharp pain in it*.*” (Adult female patient)*.

### Compliance to pre-clinical guidelines

Only 70 participants (19%) complied to the guidelines and reported that they washed the wounds with water and soap and presented to a healthcare facility within 48 hours. Of these, 45% (32/70) applied an antiseptic in addition to washing. However, 19/376 (5%) washed with water only and 183/376 (48.7%) neither washed the wound not applied anything. Overall, the commonest material applied on the wound by the 193 victims conducting pre-clinical care were antiseptic (46), herbs (25) black stone (10) unknown creams or other materials such as beans, dog urine, dust, tobacco, coins, brake fluid, acid, powder made out of dog hair and salt. Notably only 8 out of 29 study participants who have had previous dog bite episode complied with pre-clinical guidelines. Presentation within 48 hours was mentioned by three-quarters (74.7%) of the victims. The median (IQR) time to presentation at a health facility was 18 (41) hours. **Table 4** shows that compliance differed by education status (p<0.001), employment status (p = 0.01) and accessing information about dogs (p = 0.005).

**Table 4:**
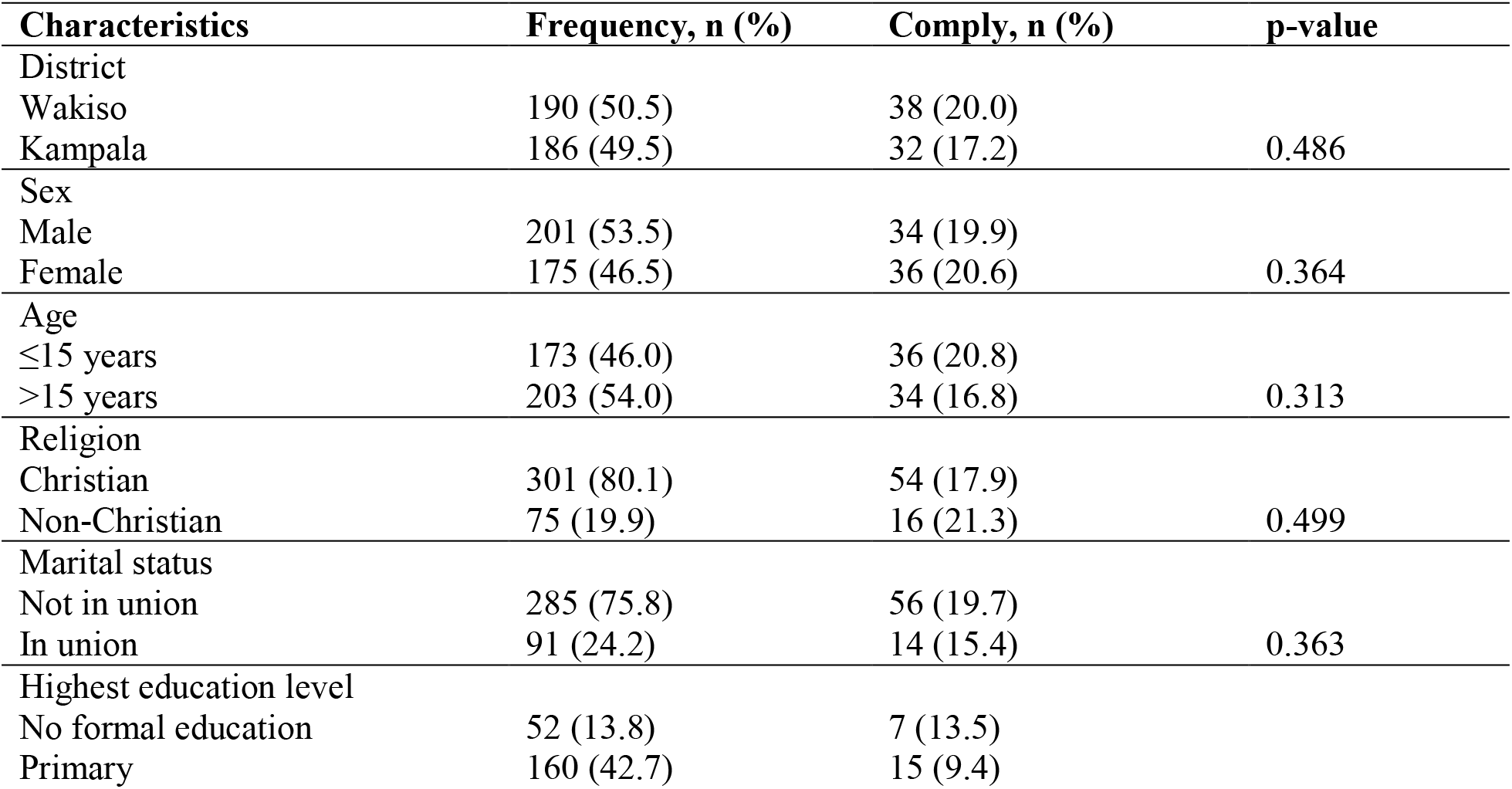
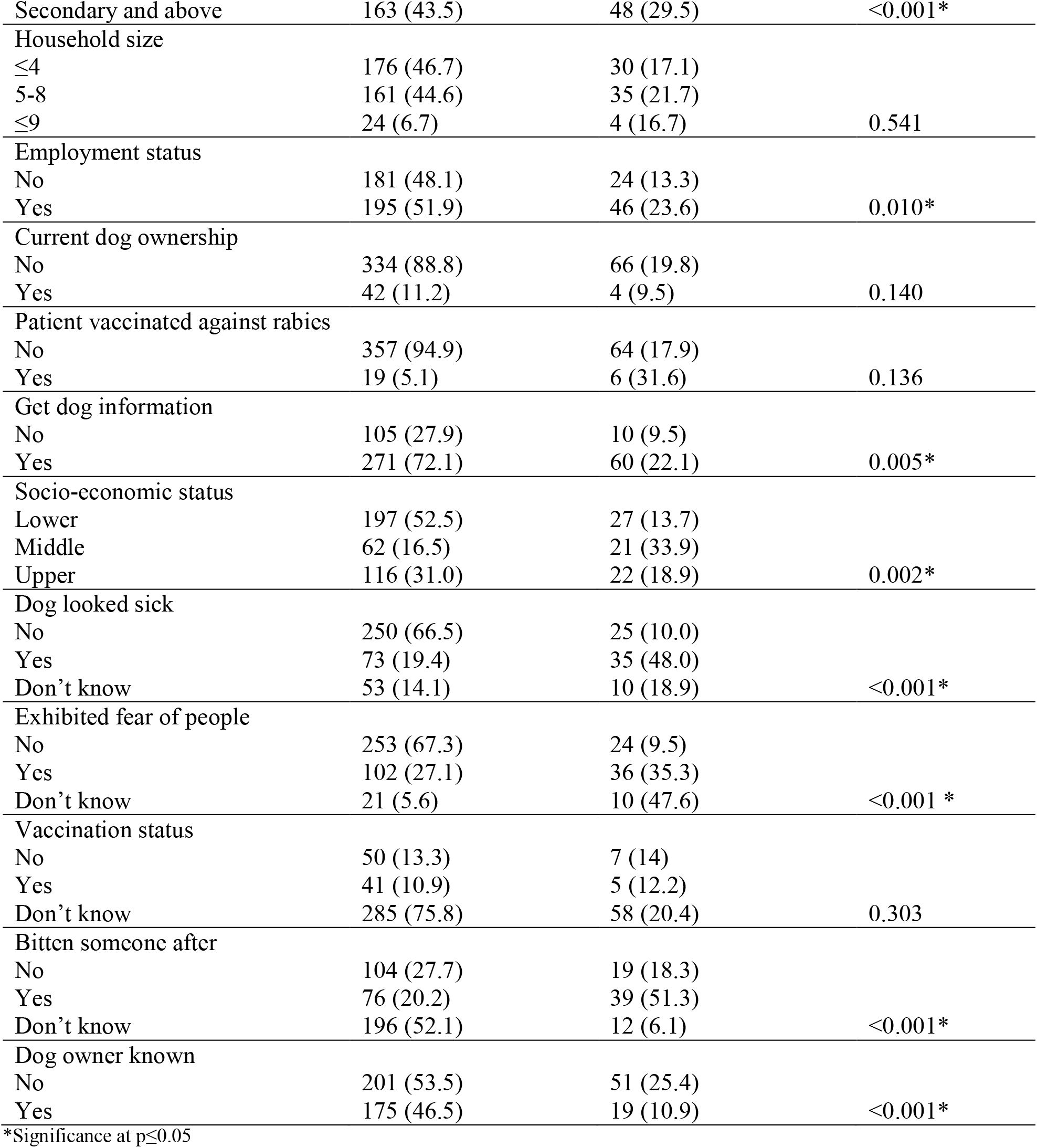
Distribution of selected characteristics of 376 respondents by compliance

### Explanations for application of non-recommended substances

#### To kill micro-organisms

On deeper inquiry, some respondents thought that by applying substances of unusual pH or temperature, they would kill the rabies virus. This came out as one of the reasons why some applied hot water, salt and battery acid as one explains;

> *“When the dog came and bit me, many of my colleagues in the garage where I work told me to first put battery acid to kill the germs* [virus] *that cause dog madness* [rabies] *before they could go very far inside the meat* [flesh]. *So, they removed the battery from the car and drained its acid into the wound here* [shows hand].*” (Male, adult patient)*.

#### Routine management of wounds

Some respondents had witnessed routine wound management with certain substances or by some procedures. It was the reason they managed the dog bite in a similar way without the specifics of a dog bite as one explains:

> *“At times you find people with a bandage. When you ask them why, they tell you they do not want the blood to move to the brain carrying dog poison. They think rabies is like snake poison that travels in the bloodstream*.*” (local veterinarian)*.

#### Knowledgeable caretakers and trust in herbalist

Additionally, some victims did not apply herbs out of choice but relied on the knowledge, skills and practices of first responders who they thought were more knowledgeable in managing dog bite. This was more pronounced when the caretaker also doubled as the decision-maker on which line of treatment to take. Similarly, a number of respondents applied herbs because they trusted the herbalist. This trust extended to the treatment which they took without questioning as one recounts:

> *“My mother sent me to the traditional doctor* [herbalist]. *There is some powdered medicine he tells you to put under the tongue then he cuts you on the leg here like this* [shows around the ankle] *then he puts black stone He told me to go home and not to bathe using cold water drink cold drinks I did not ask, I just followed instructions, it is my mother who had sent me to him”*. *(Female adult patient)*.

#### Pedigree of herbalist

The pedigree of a particular herbalist also played a key role in informing the decisions of victims. Some respondents based their decisions on success stories they had heard as one herbalist explains:

> *“they come because I have a history of healing them since the 70s. Even when they go to Mulago* [hospital], *some pass here. People believe in me. My treatment is cheap because over time, I have found out that dogs bite the poor. They should thank God, not me, for He has kept me longer*.*” (Herbalist for dog bite victims)*.

#### Perceived high cost of conventional treatment

However, some patients sought herbalist assistance because they thought they could not afford conventional treatment. These only went to hospital when they learnt that treatment was free as one elaborates;

> *“I sent my girl* [daughter] *to the herbalist, and I did not go because I did not have money for both of us. I first felt much pity for this young one* [smiles]. *Me I stayed and put tobacco. But when the dog had died, I was worried, I went to Mulago* [hospital] *after a week where I learnt that the treatment was free. I went back home and brought my daughter too. She didn’t go back to the herbalist again*.*” (Adult female patient and mother to a patient)*.

#### Conflicting information on efficacy of both herbs and modern treatment

When we investigated why some of the patients used conventional and non-conventional medicine at the same time, they pointed to information from fellow patients they found in hospital. Another reason they gave for simultaneous resort was the conflicting information proving and disproving efficacy of herbs. Therefore, they chose to use two lines as one elaborates:

> *“I went to the herbalist because our family knew very well that he works well on dog bites one of my daughters healed well, so I was sure that his medicine* [herbs] *heal those bitten by dogs. But when our LC* [local leader] *told me that in Mulago treatment was more effective and free, I also decided to come this side* [hospital].*” (Adult female patient)*.

### Explanations for seeking medical care from hospital

#### Mistrust in herbalists

Some patients talked about the mistrust they had in herbalists, even when some of them patients first went to them. They indicated dissatisfaction with the herbalist’s procedures. Some of them deliberately refused the processes and left for hospital without applying any herbs as one narrates;

> *“Now to go to Mulago* [hospital], *it has professional doctors but the one they had directed me to is a herbalist. He even wanted to cut my leg and put black stone. He did not wear gloves, so I refused. That is why I stopped him from him adding more things on my wound. I went away” (Adult male patient)*.

#### Knowledge and experiences on dangers of dog bites

Knowledge of someone who had suffered negative consequences of dog bites attributed to inadequate medical care also came out as one of the reasons why some people immediately went to hospital. Such experiences were common among the victims as one recounts;

> *“People talk. There is also a time we were in Kikandwa* [place of birth] *and a child passed on. A dog bit him and he was taken to a* [herbalist] *and received treatment. After a period of some months that I can’t recall, a child started barking and passed on. This was last year. So I could not risk going to that man* [herbalist].*” (Female adult patient)*.

#### Community advice

However, other respondents attributed their action of seeking medical care paradoxically to both mistrust and trust in community advice. Those who mistrusted community advice questioned the efficacy of different applications that were suggested to them. However, those who trusted community advice heeded and went to the hospital.

### Factors associated with compliance to standard preclinical management guidelines for victims seeking post-exposure prophylaxis

In the adjusted analysis, factors significantly associated with higher likelihood of compliance to pre-clinical guidelines were having a formal education (adjPR = 1.76, 95% CI: 1.24 – 3.79, p= 0.024), being in employment (adjPR = 1.48, 95% CI: 1.09 – 2.31, p = 0.047), perceiving the dog as being sickly (adjPR = 1.47, 95% CI: 1.02 – 2.72, p = 0.042) and knowing that the dog went on to bite another person (adjPR = 1.69, 95% CI: 1.01 – 2.86, p = 0.048). Lower likelihood of compliance was associated with being older than 15 years of age (adjPR = 0.70, 95% CI: 0.47 - 0.92, p = 0.045), not being certain whether the dog went to bite another person or not (adjPR = 0.35, 95% CI: 0.17 - 0.70, p = 0.003) and knowing the owner of the biting dog (adjPR = 0.65, 95% CI: 0.36 - 0.93, p = 0.034). Important to note is that sex and rabies immunization status of the victim did not have any bearing on the compliance as shown in **Table 5**. Notably, the interaction effects between sex and age as well as sex and marital status on compliance were not significant.

**Table 5:** Multivariable analysis of factors associated with compliance to standard preclinical management guidelines for 376 victims seeking post-exposure prophylaxis in the 2 selected hospitals in Uganda.

| Characteristics | Unadjusted<br>PR (95% CI) | p-value | Adjusted<br>PR (95% CI) | p-value |
| --- | --- | --- | --- | --- |
| District |  |  |  |  |
| Wakiso | 1 |  |  |  |
| Kampala | 0.86 (0.56 - 1.32) | 0.488 |  |  |
| Sex |  |  |  |  |
| Male | 1 |  | 1 |  |
| Female | 1.22 (0.79 – 1.86) | 0.365 | 1.04 (0.73 – 1.49) | 0.798 |
| Age |  |  |  |  |
| ≤15 years | 1 |  | 1 |  |
| >15 years | 0.81 (0.53 - 1.23) | 0.315 | 0.70 (0.47 - 0.92) | 0.045* |
| Religion |  |  |  |  |
| Christian | 1 |  |  |  |
| Non-Christian | 1.19 (0.72- 1.96) | 0.495 |  |  |
| Marital status |  |  |  |  |
| Not in union | 1 |  |  |  |
| In union | 0.74 (0.39 - 1.41) | 0.364 |  |  |
| Highest education level |  |  |  |  |
| No formal education | 1 |  | 1 |  |
| Primary | 0.70 (0.30 - 1.62) | 0.400 | 0.89 (0.81 – 2.05) | 0.783 |
| Secondary and above | 2.19 (1.05 – 4.54) | 0.036 | 1.76 (1.24 – 3.79) | 0.024* |
| Employment status |  |  |  |  |
| No | 1 |  |  |  |
| Yes | 1.78 (1.13 – 2.79) | 0.012 | 1.48 (1.09 – 2.31) | 0.047* |
| Current dog ownership |  |  |  |  |
| No | 1 |  |  |  |
| Yes | 0.48 (0.18 - 1.25) | 0.135 |  |  |
| Immunised against rabies |  |  |  |  |
| No | 1 |  | 1 |  |
| Yes | 1.76 (0.88 – 3.54) | 0.112 | 1.48 (0.81 - 2.74) | 0.203 |
| Get dog information |  |  |  |  |
| No | 1 |  | 1 |  |
| Yes | 2.32 (1.23 – 4.37) | 0.009 | 1.40 (0.74 - 2.66) | 0.295 |
| Socio-economic status |  |  |  |  |
| Lower | 1 |  |  |  |
| Middle | 2.47 (1.51 – 4.05) | <0.001 | 1.29 (0.82 - 2.05) | 0.269 |
| Upper | 1.38 (0.83 - 2.31) | 0.216 | 1.01 (0.63 – 1.62) | 0.292 |
| Perceived health status of dog |  |  |  |  |
| Healthy | 1 |  |  |  |
| Sickly | 4.79 (3.08 - 7.46) | <0.001 | 1.47 (1.02 – 2.72) | 0.042* |
| Don't know | 1.89 (0.96 - 3.69) | 0.064 | 1.29 (0.63 – 2.45) | 0.430 |
| Exhibited fear of people |  |  |  |  |
| No | 1 |  |  |  |
| Yes | 3.72 (2.34 - 5.91) | <0.001 | 1.53 (0.88 – 2.67) | 0.132 |
| Don't know | 5.01 (2.79 – 9.05) | <0.001 | 1.52 (0.79 – 2.91) | 0.931 |
| Rabies vaccination status |  |  |  |  |
| of dog |  |  |  |  |
| No | 1 |  | 1 |  |
| Yes | 0.87 (0.30 - 2.54) | 0.801 | 0.72 (0.28 - 1.83) | 0.491 |
| Don't know | 1.45 (0.70 - 3.00) | 0.312 | 0.96 (0.38 - 2.45) | 0.931 |
| Bitten someone after |  |  |  |  |
| No | 1 |  |  |  |
| Yes | 2.81 (1.77 - 4.46) | <0.001 | 1.69 (1.01 - 2.86) | 0.048* |
| Don't know / not certain | 0.34 (0.17 - 0.66) | 0.002 | 0.35 (0.17 - 0.70) | 0.003* |
| Dog owner known |  |  |  |  |
| No | 1 |  |  |  |
| Yes | 0.43 (0.26 - 0.69) | 0.001 | 0.65 (0.36 - 0.93) | 0.034* |

\*Significance at p-value $\leq 0.05$ and 95% confidence interval (95% CI).

## Discussion

The study investigated the epidemiology of dog bites and preclinical practices for the victims in the context of dog bite prevention and rabies prevention respectively. The finding that there were more males than females is in concurrence with majority of studies that have reported a preponderance for males [9, 22]. Some authors attribute this to personality variation between genders with more males being subject to dog bites [23]. Others have attributes it to males being frequently involved in day and night activities [9]. However, our findings contradict some studies which reported that females are more likely to be bitten [24].

Regardless of age, the leg was the most affected part of the body followed by hands and arms. Limbs have been documented to be the most bitten parts [25-27]. This may be attributed to accessibility, especially for the legs, and the struggles that usually ensue during the bite. Such scuffles usually involve use of arms and legs to ward off the dog. However, bites on the head were among children only and this may be explained by their height which puts the head near the mouth of the dog. Likewise, some authors have attributed this to the small physique of children, their inclination to put their faces close to animals, and limited motor skills to provide defense [22].

Majority of wounds were Category II involving skin scratches. This is expected especially when the majority of wounds were singular in number and extremities were the most affected parts. These parts are not only accessible by dogs but they are easily movable in self-defense. Given that most of the victims were walking, it was unlikely that biting dogs got a firm grasp of the victim before disentanglement. Besides, dog attacks usually last a very brief duration, which explains why very severe and fatal bites are not a common finding in literature, just like in our study. Such findings on severity are consistent with other studies [28] though they conflict with some [29].

The owner of the biting dogs was not known in most cases (53.5%). This is perhaps because majority of victims were bitten on the road or in public places like markets. Notably, the study area is mostly urban, and characterized by rapid urbanization, high population of people, abundance of garbage heaps that serve as a source food for dogs and un-owned animals especially dogs. In addition, it might be due to some dog owners not chaining their dogs and leaving them to wander hence posing a risk of bites to strangers. Some authors have attributed it to weak legislation on responsible dog ownership [30]. Just like in our study, the increased risk of bite events by such dogs compared to those with known owners has been reported in India [31] and Nigeria [26] though in Mozambique [32], they play a minor role. This shows that the role of wandering dogs in the bites may vary with each setting.

For majority of bites, it is the victims that approached the dogs rather than the other way round. Territorial invasion easily forces dogs to bite out of self-defense. Such a risk increases when dogs are in a pack or nursing young ones as explained in the in-depth interviews for our study. Studies have widely reported increased dog aggression due to territorial invasion especially by children [33, 34]. Our findings on this are consistent with that of related study in the United Kingdom which reported 50% of the victims as having approached the biting dog [35].

Before presentation to hospital, only 18.6% of the patients had complied with recommended preclinical guidelines. The WHO recommends meticulously flushing of wounds with water and soap and application of an antiseptic like povidone iodine if available [2]. The low level of compliance in our study may be due to inadequate knowledge on the guidelines. Moreover, many respondents expressed not knowing what to do immediately the dog bit them. However, our prevalence is comparable with that reported in India which varied between 2 −21% depending on the township [36]. Nonetheless, in India still, another study reported a higher rate (58%) than ours though there was a significant rural-urban divide with the former performing worse [37]. This, combined with an 7% - 45% prevalence of wound washing with soap and water in Kenya and Ethiopia respectively [38, 39], is evidence of how the practice varies across communities. It may also indicate variations in the coverage and uptake of health education interventions across societies.

Of those who applied some substances before reporting for PET, only 23.8% applied an antiseptic as recommended. A comparable proportion applied herbs whereas others used antibiotics, black stone, charcoal, acid, powder made by burning hair of biting dog, split beans, paraffin, salt, monetary coins and others. The inquiry into application of non-recommended material revealed that such practices were driven by a number of factors including individual beliefs on efficacy, lack of funds to pay for medical services as well community influences and advice. Such practices have been reported elsewhere with higher magnitudes being reported in both community [36, 37] and hospital based surveys [40].

Victims who were bitten by a dog with known ownership were 35% less likely to comply. Sometimes it is intuitive that a person bitten by a dog of known ownership might be more confident with regard to the health status of the dog compared to a bite by a dog they do not know. If the owner is known, it is easier to inquire about the health aspects of the dog like the rabies vaccination status. However, this practice of victims assessing the risk of rabies to be low based on knowing the dog ownership is dangerous and should be discouraged. Nonetheless, our findings are in concurrence with another study in Ethiopia which found that the likelihood of the dog bite victim visiting a healthcare facility more than doubled when the victim was bitten by a dog of unknown ownership [41].

Victims who were employed at the time of the bite were approximately one and half times more likely to comply with the guidelines than those who were not. This may probably be due to the fact that the employed tend to have higher education levels. Besides, employment has been associated with appropriate health seeking behavior in some studies [42]. Similar, those who perceived the biting dog as being sick were more than twice likely to comply compared to those who perceived them as being healthy. It may be that victims attached the sickness perception to an elevated risk for rabies and therefore complied to the pre-clinical preventive measures. Besides, some studies have described the health status of a biting animal as a drive to PET compliance [43]

Participants who had attained at least secondary education or higher were more likely to comply with pre-clinical guidelines compared to those with no formal education. People who are more educated tend to have a higher ability of interpreting health education messages. Moreover, in our study, those with secondary education or more were more likely to access information on dogs and dog bites than those with less education level. Our findings and probable explanation are coherent with research that has suggested that people with higher education tend to have more knowledge about rabies than illiterate ones [44, 45].

Patients aged above fifteen years were less likely to comply compared to those below 15 years. this finding is in consistent with that of a study in China that found age ?15 as being at more risk of failure to begin PEP [46]. Prioritization of younger ones to receive healthcare was evident in this study. When asked why the daughter was sent to receive treatment and the mother stayed home yet they had both been bitten by the same dog, the mother explained that the young one had more need for treatment. So, with limited resources and lack of knowledge that treatment was free, priority seemed to be given to younger ones.

Patients who did not know whether the biting dog had gone on to bite other people were 65% less likely to comply. Cases of a single dog being responsible for multiple bite cases have been widely described [47, 48] and this is typical of wandering dogs. The finding that some people did not know may be a reflection of the care-free attitude of such individuals towards the risk of bite consequences. Not caring to find out whether the dog bit other people makes them not likely to comply as they may not know the value in ascertaining the status of the dog. Nonetheless, deeper inquiries revealed that some respondents did not comply even after knowing that the dog had gone on to bite other people. However, they attributed this to lack of funds to seek treatment.

The main limitation of this study may be the self-reports about the events which might have introduced recall bias through inaccuracies in detailing the events. However, we verified the information where possible by triangulation. In addition, we used a hospital-based convenience sample and this limits the representativeness of our results for the entire population of dog bite victims. An example of this may be that there may be specific factors that influenced our respondents to report to hospital but not those who stayed home and used domestic remedies. Therefore, the findings should be interpreted within this context.

## Conclusions

The study presents evidence to show that dog bites in the study area are widespread across gender and age. The bites are both provoked and unprovoked and are majorly by wandering dogs in public places like roads. Compliance to recommended pre-clinical guidelines is low and mainly due to inadequate awareness about dangers of alternative treatments and availability of therapy. There is therefore need for holistic targeted health education programs and regulation of herbalist activities. In addition, approaches that reduce human-dog interactions in public places for example reduction of stray dog populations, need emphasis.

## Data Availability

The data that support the findings of this study are available on request from the corresponding author, SK. The data are not publicly available because they contain information that could compromise the privacy of research participants.

## Acknowledgements

We are grateful to the respondents who participated in this study and the staff of Mulago National Referral Hospital and Entebbe General Referral Hospital for providing a conducive environment for successful data collection.

This research was supported by the Consortium for Advanced Research Training in Africa (CARTA). CARTA is jointly led by the African Population and Health Research Center and the University of the Witwatersrand and funded by the Carnegie Corporation of New York (Grant No- -B 8606.R02), Sida (Grant No:54100113), the DELTAS Africa Initiative (Grant No: 107768/Z/15/Z) and Deutscher Akademischer Austauschdienst (DAAD). The DELTAS Africa Initiative is an independent funding scheme of the African Academy of Sciences (AAS)’s Alliance for Accelerating Excellence in Science in Africa (AESA) and supported by the New Partnership for Africa’s Development Planning and Coordinating Agency (NEPAD Agency) with funding from the Wellcome Trust (UK) and the UK government. The statements made and views expressed are solely the responsibility of the Fellow.

## Supporting information

**S1 Fig. Map showing the location and geographical details of the two districts from which the study participants were got**. Kampala Capital City Authority serves as the capital city of Uganda. It is divided into five administrative units called “divisions” and has an estimated human population of 1.5 million. Approximately 8% of households in Kampala own dogs. The average number of dogs owned per household is 1.7. Wakiso district is divided into 18 administrative units called sub-counties with a population of approximately 2 million people. Approximately 14% of households in Wakiso own dogs. The ownership stands at 1.9 dogs per household. However, the population of stray and free-roaming dogs in both districts is not known. Much as Mulago and Entebbe Hospitals are located in Kampala and Wakiso respectively, dog bite patients from either district can report to any of the healthcare facilities to receive PET.

**S1 Table. Variables that were studied, indicating their categorization and measures**. The factors were organized into host / patient factors, including socio-demographics and those that influence the vulnerability of people to dog bites. Some factors on biting dogs were also studied to give a clear indication of how they influence the dogs to bite as well as the practices of the victims after the bite. Factors on the circumstances of the particular dog bite event were categorized into pre-bite, during the bite and post-bite factors. The intention of this was to study why the event happened and the wound management practices thereafter. The measures indicate how the variable was recorded and / or categorized.

**S2 Table. Characteristics of biting dogs as reported by the 376 study participants from Wakiso and Kampala districts, Uganda**. The characteristics were reported by the dog bite victims or their caretakers, if they knew the details of the biting dog. The frequencies of such characteristics are presented by the specific district of residence and the differences in distribution of characteristics is indicated by the corresponding *p-value*. Notably, all study participants were residents of the district where the bite event happened.

**S3 Table. Summary of circumstances of the dog bites, immediate actions taken by victims and reasons for different applications and health seeking behavior**. Ten themes were synthesized out of the in-depth interviews to explain the circumstances in which bite events happened. Immediate actions taken by bite victims were categorized into four. For those who applied different substances to the bite wounds, the reasons for their choice and actions were recorded into five categories. The same was done to explain why victims went to herbalists, healthcare facilities or had a simultaneous resort.

